# Knowledge mobilisation of rapid evidence reviews to inform health and social care policy and practice in a public health emergency: appraisal of the Wales COVID-19 Evidence Centre processes and impact, 2021-23

**DOI:** 10.1101/2023.11.30.23299238

**Authors:** Micaela Gal, Alison Cooper, Natalie Joseph-Williams, Elizabeth Doe, Ruth Lewis, Rebecca Jane Law, Sally Anstey, Nathan Davies, Amy Walters, Robert Orford, Brendan Collins, Lisa Trigg, Chris Roberts, Sarah Meredith, Steven Macey, Andrew Carson-Stevens, Jane Greenwell, Ffion Coomber, Adrian Edwards

## Abstract

The Wales COVID-19 Evidence Centre (WCEC) was established from 2021-23 to ensure that the latest coronavirus (COVID-19) relevant research evidence was readily available to inform health and social care policy and practice decision-makers. Although decisions need to be evidence-based, ensuring that accessible and relevant research evidence is available to decision-makers is challenging, especially in a rapidly evolving pandemic environment when timeframes for decision-making are days or weeks rather than months or years. We set up knowledge mobilisation processes to bridge the gap between evidence review and informing decisions, making sure that the right information reaches the right people at the right time.

**Aims and objectives:** To describe the knowledge mobilisation processes used by the WCEC, evaluate the impact of the WCEC rapid evidence reviews, and share lessons learned.

**Methods:** Our knowledge mobilisation methods were flexible and tailored to meet stakeholders needs. They included stakeholder co-production in our rapid evidence review processes, stakeholder-informed and participatory knowledge mobilisation, wider dissemination of outputs and associated activities including public engagement, capacity building and sharing of methodologies. Feedback on processes and evidence of impact was collected via stakeholder engagement and a stakeholder survey.

**Results:** Findings indicate that WCEC knowledge mobilisation processes successfully supported co-production and use of rapid evidence review findings by scientific advisors and policy and practice decision-makers during the COVID-19 pandemic. Identified barriers and facilitators are of potential relevance to wider evidence initiatives. Knowledge mobilisation require sustained development to continue building stakeholder links, embed co-production and sustain knowledge mobilisation as we move to support evidence-based policy and practice decision-making beyond the pandemic.

**Discussion and Conclusion:** The WCEC knowledge mobilisation processes successfully supported co-production and use of rapid evidence review findings by scientific advisors and policy and practice decision-makers during the COVID-19 pandemic. Identified barriers and facilitators are of potential relevance to wider evidence initiatives. Knowledge mobilisation require sustained development to continue building stakeholder links, embed co-production and sustain knowledge mobilisation as we move to support evidence-based policy and practice decision-making beyond the pandemic.

**Funding statement:** The Wales COVID-19 Evidence Centre was funded for this work by Health and Care Research Wales on behalf of Welsh Government.

## 1.0 Introduction

The COVID-19 pandemic dramatically changed health and social care needs and the way essential services were delivered in Wales and beyond. Health and care policy and practice decisions had to be rapidly made and informed by evidence. The need for evidence-informed policy and practice decisions is undisputed.(1) However, ensuring that evidence is available and accessible is challenging at the best of times and even more so in the context of a rapidly evolving public health emergency.(2)

The National Institute of Health and Care Research (NIHR) describes knowledge mobilisation as *‘Getting the right information to the right people in the right format at the right time, so as to influence decision-making’*, and hosts a range of resources for knowledge mobilisation.(3) Knowledge mobilisation is a dynamic and iterative process that includes engagement, co-production, shared learning, dissemination, communication, and the exchange and use of knowledge. Knowledge mobilisation is integral to the research translation pathway that helps to bridge the gap between research findings and use of the findings to achieve real world benefit and impact. Several frameworks, theories, models and guides for knowledge mobilisation have been developed including the widely used and adapted ‘Knowledge to Action’ Framework, Developing Evidence Enriched Practice, the ‘What Works Networks’, and BRIDGE criteria; the latter two being targeted towards implementing research evidence in the policy context.(4–7) In 2021, Social Care Wales (a Welsh Government-Sponsored Body) published its model for knowledge mobilisation, setting out its aims and approach for evidence-enriched practice, planning and policymaking in social care in Wales.(8)

There is much published research on the enablers and barriers to the use and implementation of research evidence. In the policy and practice context, common barriers include a lack of co-production to ensure research relevance and need, inaccessibility of the evidence, and failing to meet the timeframe of the policy cycle or practice need.(1,9–11) Conversely, enablers include early engagement and co-production of research together with end users, which helps researchers to understand the setting and timeframe for evidence uptake. This ensures that research addresses a real need, and that the findings are accessible and more likely to be used. It should be noted that research evidence will only partly contribute towards the knowledge used by decision-makers, who will also use advice from domain experts, from those with lived experience, and their own knowledge to inform decisions.(12)

Achieving impact from research is enabled by knowledge mobilisation. While impact is often referred to as public and patient benefit, other forms of impact are also valid, with many being achievable along the translation pathway to patient and public benefit. Shorter term impact can include increasing knowledge or awareness, or changes in attitudes and motivation that can contribute to later changes in individual practice. Longer-term outcomes are the changes in policy, behaviour or practice that benefit patients and the public, which can take years.(13)

### 1.1 The Wales COVID-19 Evidence Centre

Several new entities were set up to conduct rapid reviews of research evidence to aid decision-makers during the COVID-19 pandemic. Examples include the UK Health Security Agency COVID-19 Rapid Evidence Service, the COVID-19 Evidence Network to support Decision-making (COVID-END Global), and the WCEC.(14–16) With the remit of ‘*Good questions, answered quickly’*, the WCEC was funded by Welsh Government and rapidly set-up (March 2021 – March 2023) to collect research evidence and ensure this was accessible and rapidly available to the people making decisions for health and social care policy and practice in Wales during the pandemic and in the later move towards recovery. The WCEC funding covered its partner research groups and public partner members but not stakeholder time or commitment which was given *gratis* throughout the work.

Knowledge mobilisation underpinned all the WCEC processes including stakeholder identification, research question prioritisation, rapid evidence reviews, and also rapid primary research and public involvement (the two latter are not included in this paper).

### 1.2 Aims

The aims of this paper are to describe, evaluate and reflect on the processes for knowledge mobilisation and evidencing impact from the WCEC rapid research evidence reviews, and make recommendations for best practice.

## 2.0 Materials and methods

Knowledge mobilisation and evidencing impact were a priority for the WCEC, with two members of staff employed to support this, and a member of the Welsh Government Technical Advisory Cell (TAC) working closely with the WCEC core team to facilitate engagement and communication between Welsh Government teams and the Centre.

Knowledge mobilisation processes were iterative, tailored to meet the requirements of stakeholders and included the following activities: 1) co-production and engagement with stakeholders, 2) stakeholder informed knowledge mobilisation, 3) wider dissemination, 4) associated knowledge mobilisation activities, and 5) tracking and evidencing impact (Fig 1). We describe each step below.

**Fig 1.**
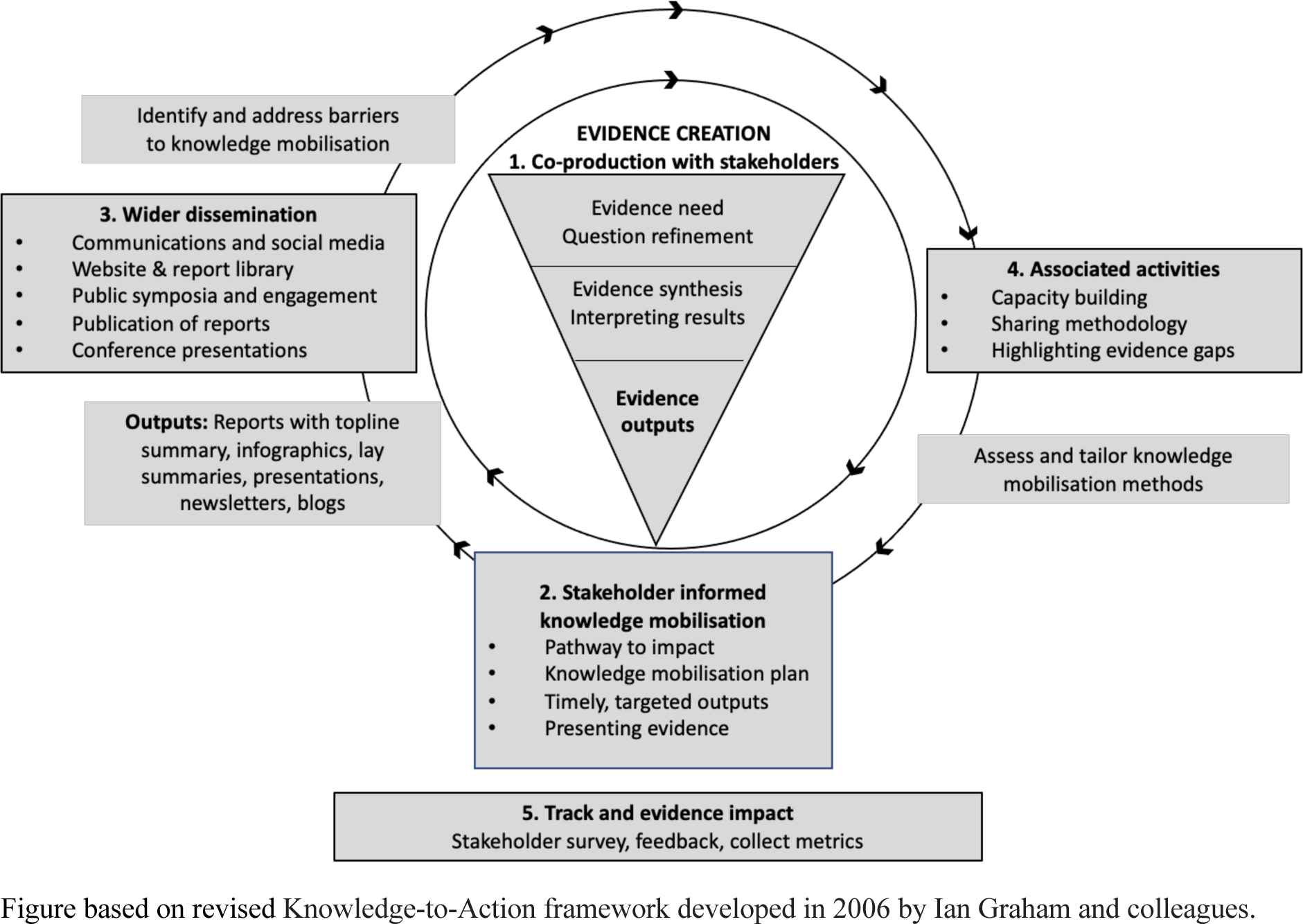
Knowledge mobilisation and impact: the 5 steps and areas of activity.

### 2.1 Co-production and engagement with stakeholders (step 1)

The WCEC identified and reached out to stakeholder groups in Wales to identify research questions of the highest priority for health and social care decision-making during the pandemic and subsequent recovery period. Stakeholders engaging with the Centre included teams from Welsh Government’s TAC and Technical Advisory Group (TAG) (providing a role for Wales akin to “UK Government Scientific Advisory Group for Emergencies (SAGE)”) described in their terms of reference,(17) which provided coordination of scientific and technical advice to support Welsh Government decision-makers during the pandemic.

Other stakeholders included TAG sub-groups (e.g., Risk Communication and Behavioural Insights, Policy Modelling, and Environment), Social Care Wales, NHS and social care leaders, public and professional societies (Table 1).

**Table 1.** Stakeholders engaging with the Wales COVID-19 Evidence Centre.

|  |
| --- |
| **Welsh Government (WG) and Senedd (Welsh Parliament)** |
| Technical Advisory Cell (TAC) and Technical Advisory Cell (TAG). TAG subgroups include the All Wales Modelling Forum, Policy Modelling, Research and Development, Socioeconomic Harms, International Intelligence, Virology and Testing, Children and Young People, Risk Communication and Behavioural Insights, Environmental Science. |
| (Membership of TAC/TAG included Welsh Government, Public Health Wales, NHS Wales and academia; experts from public health, health protection, medicine, epidemiology, modelling, technology, data science, statistics, environment, microbiology, molecular biology, immunology, genomics, risk communication and behavioural insights, physical sciences, research). |
| Other WG teams: Long COVID Task Force, Equality and Human Rights Division, Homelessness prevention, Education (including Early Childhood Education and Care and Early Years Workforce, Childcare, Play and Early Years Division, Education and Public Services Group), Rural affairs team/Rural development division, Cost of Living Expert Group, Energy and climate change group, Planned Care Improvement and Recovery, Knowledge and Analytical Services, and the Health and Social Services group more generally. |
| Senedd (Welsh Parliament) Health and Social Care Select Committee, and Senedd Cross-party Group on Long-COVID. |
| **Social Care Wales** |
| Social Care Wales's Improvement and Development team |
| Association of Directors of Social Services (Wales) |
| **National Health Service (NHS) and other organisations aligned to health** |
| Vaccine equity committee (Welsh Government and Public Health Wales) |
| Public Health Wales Virology reference laboratory |
| Public Health Wales Vaccine Preventable Disease Programme team |
| Wales Health Board Medical Directors |
| All Wales Medicines Strategy Group |
| Academy of Medical Royal Colleges Wales, Royal College of Surgeons, Royal College of General Practitioners, Royal College of Podiatrists |
| Wales Cancer Network |
| National Health Service (NHS) leads in Wales including for primary care and cancer |
| General Medical Council-UK |
| Welsh Ambulance Trust |
| NHS Wales Shared Services partnership (Engineering team), and Infection control |
| Health Education and Improvement Wales, Deans of Medical Education (Wales) |
| UK Health Security Agency |
| NHS Wales: Directors of planning and finance, Diagnostics Board |
| **Members of the public and representative groups from under-served communities in Wales** |
| Service Users for Primary and Emergency Care Research Group |
| WCEC Public Partnership Group (8 members) |
| Members of the public attending WCEC public symposium (included workshops to identify research priorities). |
| Young people (from THE Centre for Development, Evaluation, Complexity and Implementation in Public Health Improvement (DECIPHER) ALPHA group (a research advisory group of young people aged 14 to 25 in Wales) |
| Disability Wales |
| Ethnic minority and Youth Support Team (EYST) Wales |
| Housing association (Taff Housing) |

Stakeholders’ questions were prioritised, with those most urgent and likely to have an impact (e.g., inform health and social care policy or practice decisions) being taken onto the WCEC work programme. The prioritisation process was flexible and responsive to accommodating new urgent questions. The WCEC and stakeholders worked collaboratively to develop focused research questions and agree on objectives, discuss evidence review findings and relevance to the local setting (Wales), identify policy and practice implications, write and review the final reports, plan and support knowledge mobilisation, and provide evidence of impact.

In addition to e-mail communication, co-production was enabled through generally three on-line meetings with key stakeholders during the review process, presentations of findings to the stakeholder group, and involving stakeholders in dissemination of findings (e.g., Welsh Government evidence briefing sessions, and public symposia). Meetings with stakeholders included discussions on knowledge mobilisation. Feedback to improve WCEC processes was sought from stakeholders at meetings, via e-mail and through a stakeholder survey.

### 2.2 Stakeholder informed knowledge mobilisation (step 2)

#### 2.2.1 Pathway to impact

Stakeholders submitting questions to the WCEC were asked to identify the potential impact that answering a question would have (e.g., used to inform a particular policy or decision) and the timeframe that the evidence was needed by).

#### 2.2.1 Knowledge mobilisation plans

Final evidence reports were accompanied by a knowledge mobilisation plan developed with stakeholder input (Fig 2). The plans included ‘stakeholder mapping’ to identify key individuals involved in policy or practice decision-making, and further groups with an interest in the review topic. The plan identified relevant resources and dissemination methods which would be jointly supported by the WCEC team and the stakeholders e.g., Welsh Government ‘evidence briefing’ sessions and themed public-facing symposia.

**Fig 2.**
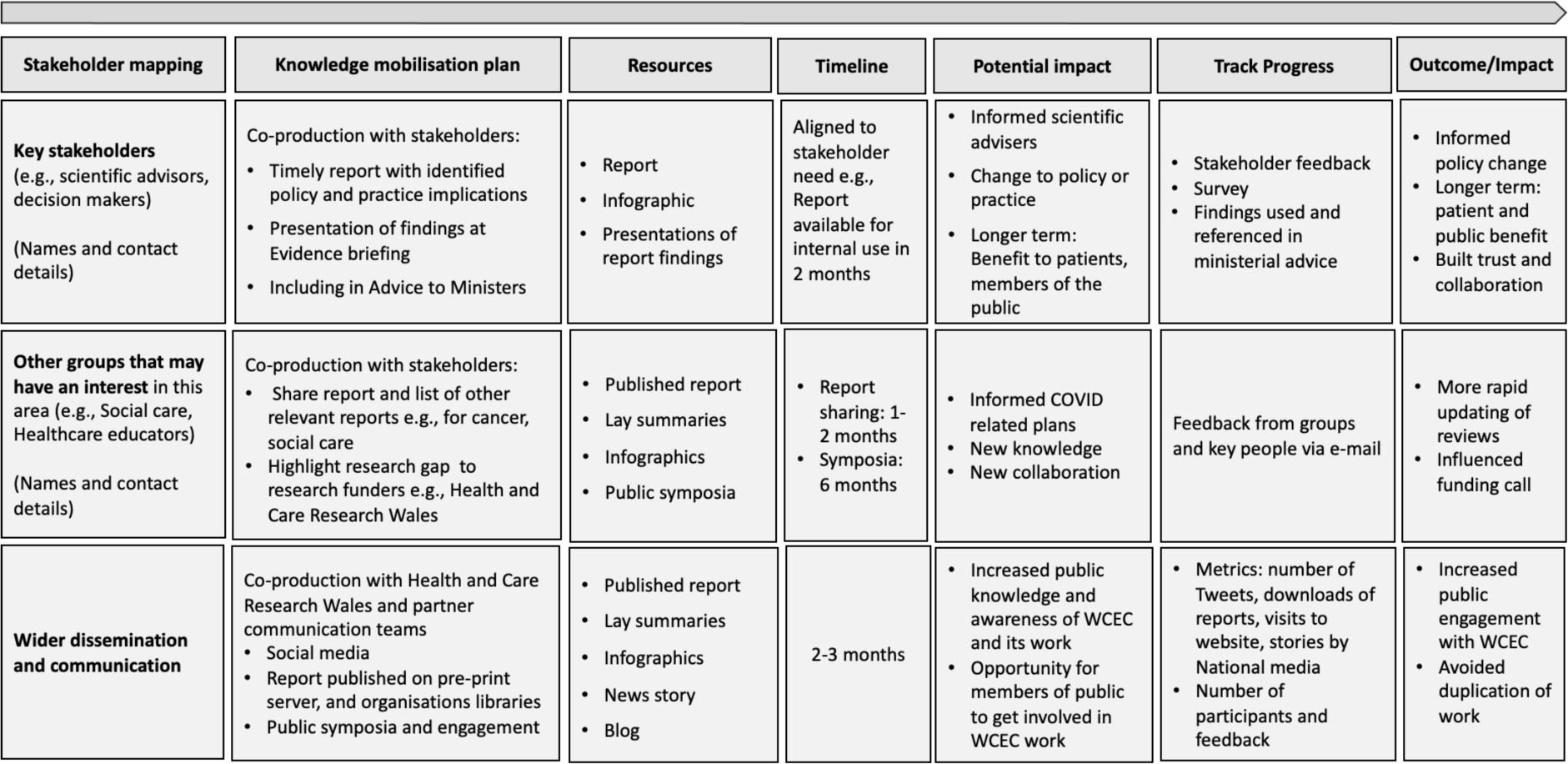
Example of a knowledge mobilisation plan.

#### 2.2.2 Timely targeted outputs

##### Reports

Evidence reviews were conducted rapidly (typically 1-3 months for rapid reviews, and from 1-2 weeks for some rapid evidence summaries. The outputs of evidence reviews were presented in reports. Report templates were developed to ensure relevant information could be easily accessed by stakeholders. Reports included a short information box relating to dates and report contributors, followed by a 2-page ‘Topline summary’ (Fig 3).

**Fig 3.**
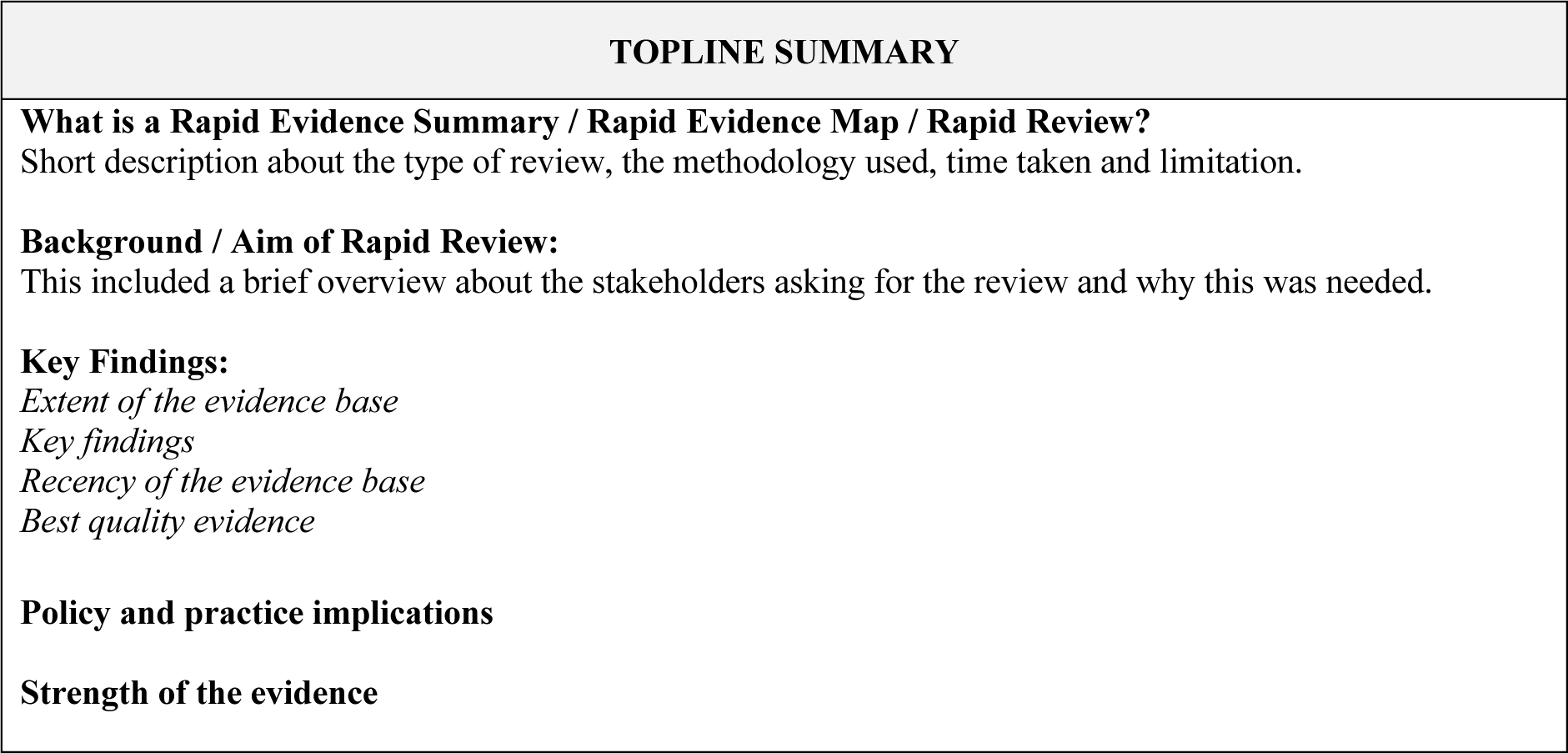
Two-page topline summary template.

Stakeholders, including the WCEC public partnership group members, contributed to report reviewing and identifying the policy and practice implications. Reports were made available to stakeholders for use within their teams at the point of completion (pre-publication) but were watermarked to limit circulation beyond those teams until publication on pre-print servers and in the WCEC library. Feedback on report structure, timeliness and accessibility of the information was collected via the stakeholder survey.

##### Infographics

When identified by stakeholders as being helpful for accessibility and dissemination purposes, a one-page infographic was produced by LD / FC using Prezi and Affinity Software. The infographics highlighted the report findings, and the policy and practice implications in a single page, using accessible text and illustrations.

#### 2.2.3 Presenting evidence

Review findings were shared via presentations to key stakeholders at the final stakeholder meetings, to the Welsh Government TAC and TAG, and at fortnightly on-line Welsh Government evidence briefing sessions. Evidence briefings were invitation-only events where questions could be posed to a panel (key stakeholders, WCEC public partnership group members, review team), and results and knowledge mobilisation discussed.

### 2.3 Wider dissemination (step 3)

#### 2.3.1 Communications and social media

Health and Care Research Wales (HCRW) developed a communications strategy (including media and social media management), provided support for the WCEC website and events, and worked to feature WCEC in the media. Communication outputs for the reports including blogs, news stories, social media and event webpages were delivered by HCRW, raising the profile of WCEC and its publications across HCRW channels. A WCEC hashtag (#WalesCovidEvidence) was used to raise social media profile. WCEC’s Public Health Wales and Health Technology Wales partner groups (NHS based) also undertook social media to raise awareness of WCEC outputs within their networks and websites.

#### 2.3.2 Website and report library

Health and Care Research Wales created, updated, and maintained an online WCEC section on their website in English and Welsh. The WCEC webpage hosted information about its work programme, methods, events, and links to news stories and WCEC newsletters.

Public facing documents including infographics, lay summaries, report Topline summaries, symposium presentations, newsletters, news stories, blogs and the WCEC work programme were translated into Welsh and hosted on the Welsh language version of the website. Full translation of reports into Welsh was available on request.

The website included an easily searchable (by theme, key words), bespoke report library. The library landing page for each report was a lay summary (written by members of the WCEC public partnership group) with a link to the full report and infographics, which could be downloaded.

A link to the WCEC website was included in the websites of other entities undertaking COVID-19 related rapid evidence reviews to raise awareness of the Centre’s work and avoid duplication. This included the UK Health Security Agency Rapid Evidence Service and COVID-END.

#### 2.3.3 Public symposia and engagement

WCEC held three themed public facing, on-line ‘Evidence into Practice Symposia’ focusing on specific areas of its work including the effects of the pandemic on education, inequalities, and public involvement. Senior Welsh Government ministers or senior policy officials or science advisors opened each of the events. The Symposia programmes included question-and-answer sessions with an expert panel (Welsh Government scientific advisors, stakeholders, and members of the evidence review teams). At one Symposium, breakout groups were included to identify questions that were important to participants, with subsequent voting to rank the top 10 priority questions for inclusion in the WCEC question prioritisation process.

WCEC took part in two public engagement events and a series of focus groups to raise awareness of the Centre and to engage and involve under-represented groups in Wales in its work. Set up, promotion and conduct of these events was supported by HCRW members. (The public engagement is not included in this paper).

#### 2.3.4 Publication of reports

Initially, completed evidence review reports were published only in the WCEC library. From May 2022, reports were published on pre-print servers including MedRxiv (health and social care relevant reports), EdArXiv (education relevant reports), and Research Square (environment relevant report). The WCEC library linked to the reports on the pre-print servers.

#### 2.3.5 Conference presentations

Members of the WCEC, including partners, were encouraged to present methods and review findings at conferences to increase visibility of the Centre, its expertise and work to academic and public health audiences.

### 2.4 Associated activities (step 4)

Associated knowledge mobilisation activities included capacity building, sharing methodologies, and identifying evidence gaps. Evidence gaps and need for further research identified during the review processes were collated with an aim to share with research funders (e.g., HCRW and the National Institute of Health Research). COVID-19 related systematic reviews and preparation of infographics were offered as projects to medical students at Cardiff University. As the timeline of student projects would not have met the rapid turnaround required for many reviews, the questions they answered were not as urgent. Stakeholders still received timely findings via a report and presentation, and students subsequently submitted to the University and peer reviewed journals.

### 2.5 Track and evidence impact (step 5)

#### 2.5.1 Stakeholder survey

A stakeholder survey was developed to capture information and feedback including on WCEC processes, report structure, ease of information access, further suggestions for knowledge mobilisation, and using the review evidence. The survey also included open ended elements to collect stakeholder feedback. Stakeholders completed the survey following the report publication, and up to three reminders were sent. Where stakeholders were involved in more than one evidence review, they were invited to include more than one report in their survey completion.

#### 2.5.2 Feedback and collection of metrics

Only questions with a clear pathway-to-impact (e.g., inform Welsh Government advisors, policy or plan) were accepted onto the work programme. Where stakeholder surveys were not completed, information pertaining to impact and use of the report was collected via e-mail, in on-line stakeholder meetings and evidence briefing sessions.

Reference of reports in stakeholder publications (e.g., Welsh Government TAC/TAG publications) was identified and collected. Reference to WCEC reports in Welsh Government publications was enabled by having a boundary-spanning Welsh Government team member in the core group to highlight the Centre’s activities within Welsh Government.

## 3.0 Results

### 3.1 Co-production and engagement with stakeholders (step 1)

No stakeholders declined to participate in WCEC’s work owing to lack of direct funding throughout its 2021-23 programme. There was good engagement from all stakeholders in WCEC reviews including i) refining the research question (usually ‘PICO’ format(18)) and agreeing relevant objectives, ii) agreeing deadlines for the evidence outputs, iii) interpreting review findings, including policy and practice implications, iv) providing input to knowledge mobilisation plans, v) supporting dissemination (e.g., evidence briefings and public symposia) and vi) providing information on how the research evidence was used.

Results from the stakeholder survey (n=21) indicated that 19 (90.5%) stakeholders were ‘very satisfied’ with the engagement process and meetings, and two (9.5%) were ‘satisfied’. (Details on survey sample and responses can be found below in section 3.5). Stakeholders reported value in being part of the review process, committed their time to attend the meetings and understood and trusted the review methods and findings:

> *‘…the WCEC representatives involved with the project were highly responsive. Their suggestions were constructive and they worked proactively to make meaningful progress that allowed our Welsh Government project team to quickly and conveniently locate relevant evidence that helped shape our policy proposals’* (S13).

Whilst positive towards the general approach, feedback suggested refining down-stream question prioritisation processes, so that the evidence review processes could be streamlined:

> *‘….I think that …. there is no perfect solution or approach. There is an evident need to manage demand and expectation at every stage of the process if the WCEC is not to be over-whelmed. there is … a need to be more rigorous (even brutal) in requiring that questions are specific and narrow (and thus deliverable) from an earlier stage but also understand that this is not easy’* (S10).

### 3.2 Stakeholder informed knowledge mobilisation (step 2)

#### 3.2.1 Knowledge mobilisation plans

Gathering information to support a knowledge mobilisation plan for each review report worked well during discussions at stakeholder meetings, evidence briefings and via e-mail follow-up. Additional information was also gathered from the stakeholder survey. Stakeholders shared the reports within their teams and in some cases also supported knowledge mobilisation more widely. For example, stakeholders from Social Care Wales arranged for findings from a rapid review of ‘Innovations help to attract, recruit and retain social care workers within the UK context’ to be presented at the 2022 Association of Directors of Social Services (ADSS Cymru) summer seminar and at a meeting of theADSS Cymru Workforce Leadership Group. They also shared the report with the Social Care Wales workforce task and finish group (community capacity building), and with regional workforce boards (Directors of Social Services, workforce leads, managers and staff in Wales).

A further example is for a review conducted on ‘the effectiveness of community diagnostic centres. Stakeholders including the Welsh Government lead for planned care improvement and recovery enabled the findings to be presented to relevant decision-making groups including at a TAG meeting and at a Welsh Government Health Executive meeting (NHS (Health Boards) Directors of Planning, and Diagnostics leads).

#### 3.2.2 Outputs and presentations

The results from the stakeholder informed knowledge mobilisation activities are presented in Table 2.

**Table 2.** Results from the stakeholder informed knowledge mobilisation activities.

| Activity | Results and feedback |
| --- | --- |
| **Reviews conducted and reports** | From April 2021 to March 2023, the WCEC conducted 51 evidence reviews and published 51 reports (in the WCEC library and on pre-print servers), which met stakeholders timelines (usually 1 week to 3 months).<br>From 21 survey responses about the report structure and accessibility, 100% found the ‘Topline summary’ the most useful part of the report. The ‘strength of evidence’ section was deemed useful by over 90% of respondents, with several commenting on the importance of this area.<br>*The highlighting of the strength of the evidence was excellent and particularly helpful. We often get evidence presented as gospel, it was good to see a more honest approach (S18).*<br>Respondents suggested improving the reports for language accessibility and highlighting of key points:<br>*‘... Within full report the key findings potentially could be more clearly presented. If bold highlighting key text, then useful to have key words rather than whole bulleted statements for example’ (S12).* |
| **Infographics** | 14 infographics were produced (example in Fig 4). Infographics were deemed a useful addition to support wider accessibility.<br>Feedback for an infographic on the ‘Effectiveness of community diagnostic centres’ included: *‘I think it’s a really useful infographic for NHS Wales .... At a time when the* |
|  | *system is so busy that Executives in particular may not initially have time to read the full evidence, the key points ....will certainly spark their interest'* (E1).<br>An example of infographic use: The review findings and infographic for 'the evidence of direct harm from COVID-19 infection and COVID-19 vaccine in pregnant/post-partum women and the unborn child' were presented at a meeting of the midwives and maternity service leads in Wales. The infographic was deemed useful to support health professionals in their discussions with pregnant women regarding COVID-19 vaccination, and the group provided feedback to improve the readability. The infographic and report were promoted to midwives, to discuss with women, via the Public Health Wales COVID-19 immunisation programme page. |
| **Presentation of review findings** | The results of 51 evidence reviews were presented to the key stakeholders at final stakeholder meetings.<br>24 Welsh Government evidence briefing sessions were held (October 2021 – January 2023), with an average of 33 participants. Feedback for the evidence briefings was positive:<br>*'Both the evidence briefings and symposium were very insightful events that provided useful opportunities to share learning and develop links for future collaboration'* (S18). |

**Fig 4.**
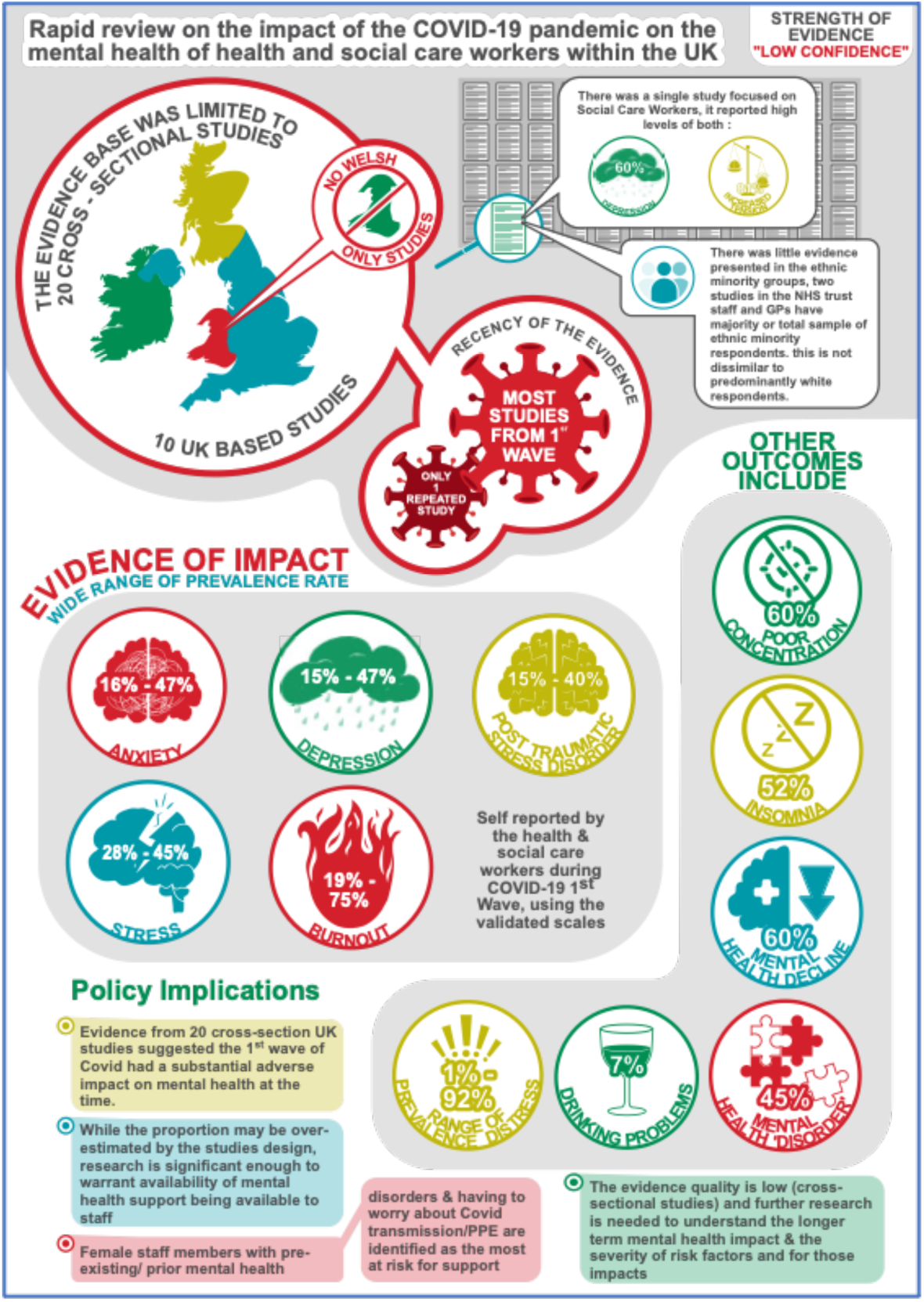
Example of an infographic produced using Affinity Designer and Publisher.

### 3.3 Wider dissemination (Step 3)

The results of wider dissemination activities are shown in Table 3.

**Table 3.**
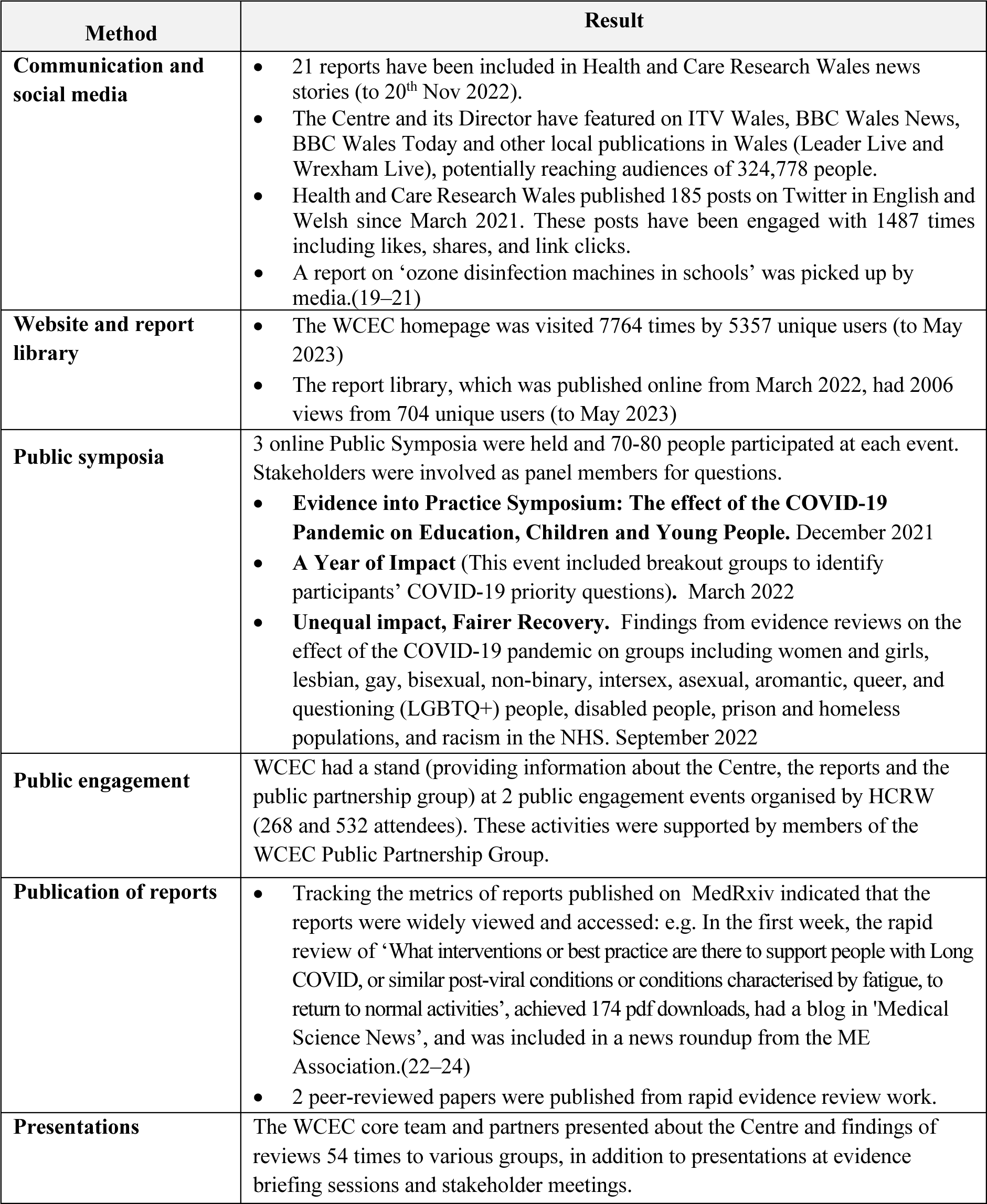
Results of wider dissemination activities.

### 3.4 Associated activities (step 4)

Four Cardiff University undergraduate medical students conducted COVID-19 systematic reviews (racism in the NHS, interventions to mitigate against racism in the NHS, impact on prison and homeless populations), and involved stakeholders to ensure outcomes were useful and relevant to inform practice. They also participated in knowledge mobilisation activities, presenting findings at Welsh Government evidence briefing sessions and public symposia. These systematic reviews have been submitted for peer-reviewed publication. 3 other students completed infographics to support WCEC evidence review reports.

Evidence gaps and need for further research that were identified in the review process were collated and are now being prioritised and formalised to share with research funders, including HCRW and the (UK) National Institute for Health Research in 2023.

WCEC shared its methodology and work programme to raise awareness of the Centre’s work and avoid duplication. This included sharing with the UK Health Security Agency Rapid Evidence Service, National Institute of Clinical Excellence (NICE) Wales, and COVID-END. The September 2021 work programme was also referenced in a Welsh Government Technical Advisory Cell (TAC) summary of advice.(25) Summaries of individual WCEC reviews were regularly included in the TAC summaries of advice.

### 3.5 Track and evidence impact (step 5)

#### 3.5.1 Stakeholder survey

All stakeholders who participated in the reviews and stakeholder meetings (1-3 per evidence review) were invited to complete the survey. However, stakeholders were variably and overall less engaged in the survey processes. Twenty-one stakeholders (mainly from Welsh Government) completed the survey (to December 2022) and two of these completed the survey for more than one evidence review. These included representatives from Welsh Government (policy lead and scientific advisors), Public Health Wales, the NHS and social care. Not all stakeholders answered every question (they were not necessarily involved in all the processes). Some respondents dropped off towards the end of the survey. Nevertheless, some useful information was obtained regarding impact, and overall satisfaction with Centre processes including those aligned to knowledge mobilisation was high (Table 4), with feedback including:

> ‘*I have seen excellent teamwork and a strong professional ethic that holds itself to account to provide the highest quality research and analysis’…. ‘The model for delivery is one that is agile and flexible as well as speedy and there is a demand for this - albeit with an understanding of the caveats that come along with it’* (S6).

**Table 4.** Survey responses for questions related to knowledge mobilisation.

| Question | Option 1<br>Number (%) | Option 2<br>Number (%) | Other options<br>Number (%) |
| --- | --- | --- | --- |
| Stakeholder engagement process (n=21) | Highly satisfied<br>19 (90.5%) | Satisfied<br>2 (9.5%) |  |
| Timeliness of the report and information (n=21) | Highly satisfied<br>18 (85.7%) | Satisfied<br>3 (8.3%) |  |
| I trusted the information in the report (n=22) | Strongly agree<br>20 (90.9%) | Agree<br>2 (9.1%) |  |
| The report presented required information in a clear manner (n=22) | Strongly agree<br>13 (59.1%) | Agree<br>8 (36.4%) | Neither agree nor disagree<br>1 (4.5%) |

There were no overtly critical responses, and a general consensus that the approaches used worked well.

A suggestion for wider dissemination of findings in other engaging formats was made (e.g., review findings relevant to children and young people could be made into a poem). This was not possible at the time due to the large number of reports and constraints during the pandemic.

#### 3.5.2 Feedback and collection of metrics

The evidence reviews informed scientific advisors, health and social care policy and practice decision-makers, and their teams. Where the reviews identified a lack of published evidence, or that evidence was of low quality, the reports indicated the need for robust research and evaluation. In some cases, there was little evidence available in particular areas, and sometimes there was no further information than that which had already been identified by stakeholders:

> Whilst the evidence of proven innovations was quite weak some of the evidence provided mirrored a lot of the initiatives that are already underway in Wales. The next phase is to further understand what practice is being adopted operationally and we have plans to do just that’ (S14).

Twenty-one review reports were referenced in Welsh Government advice and reports, and informed advisors and policy and practice decisions. Examples are provided in Table 5.

**Table 5.**
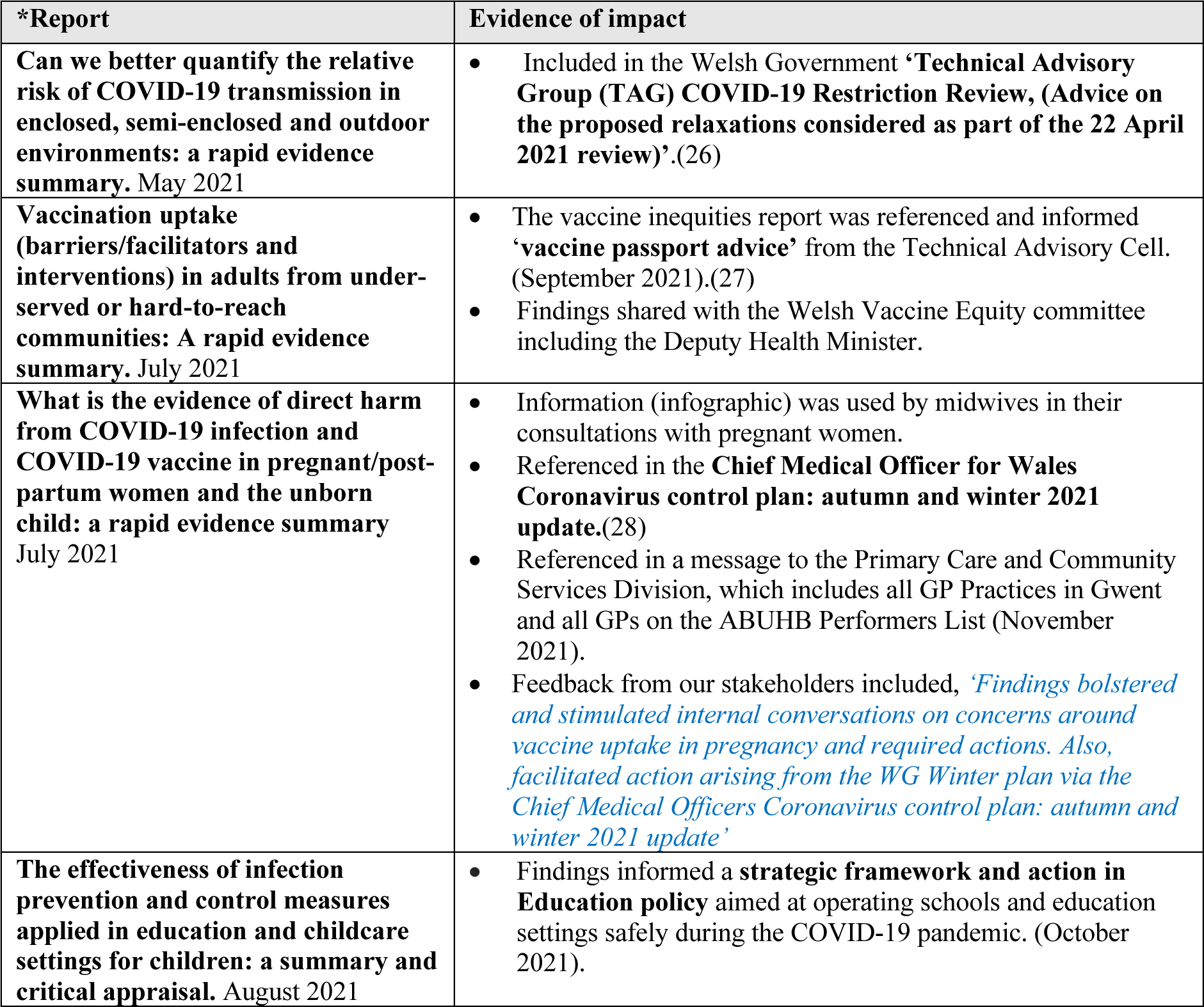
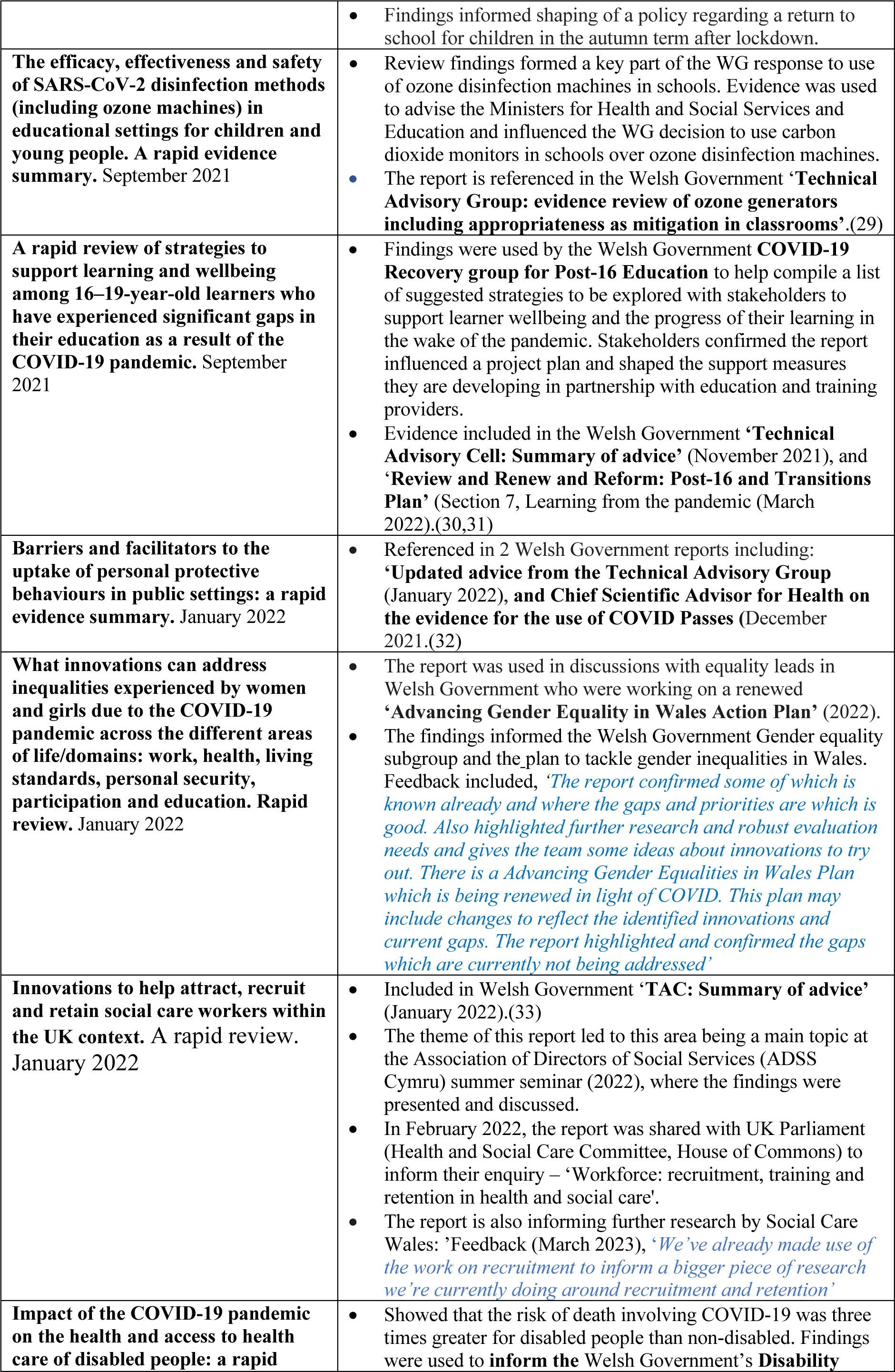
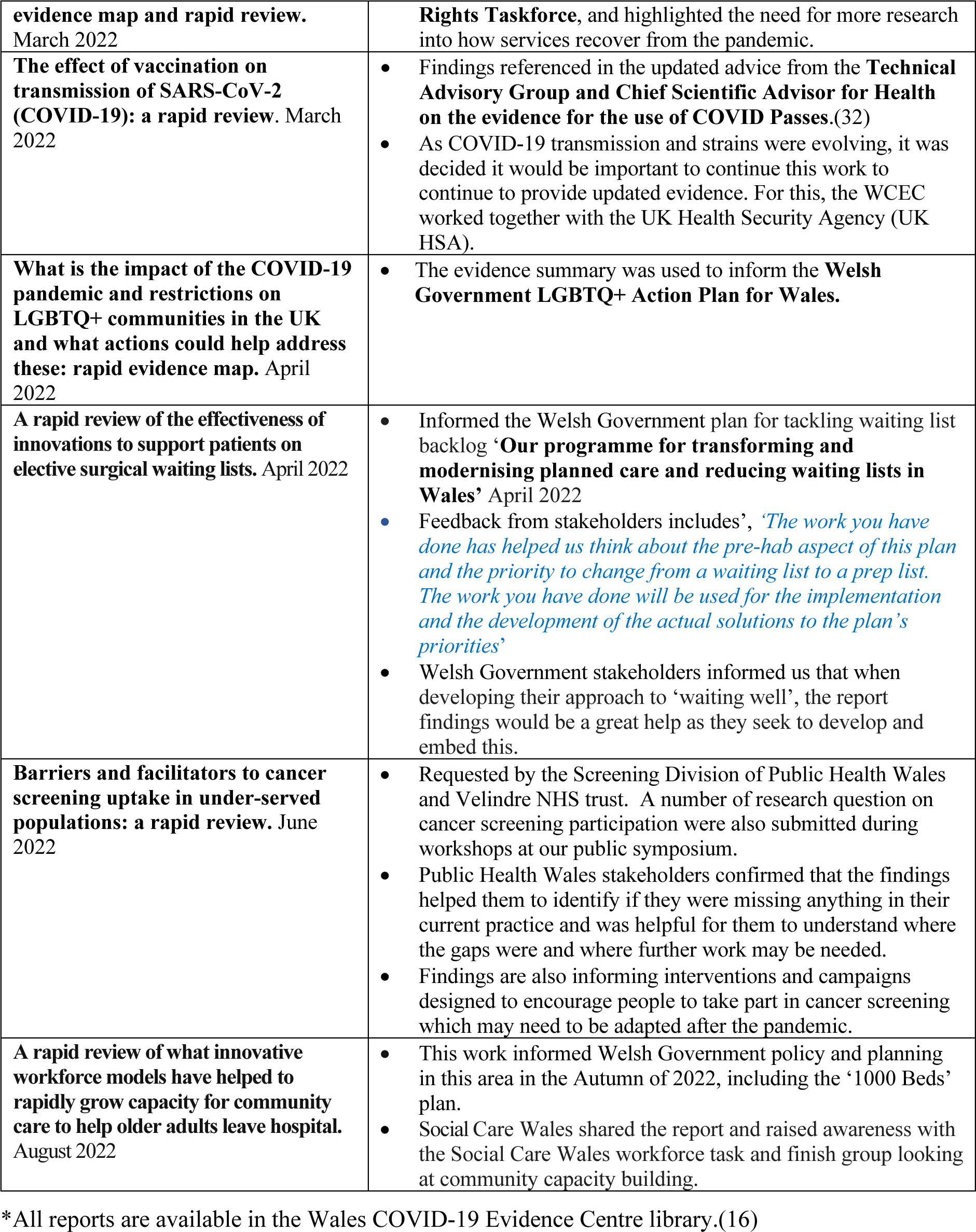
Examples of WCEC rapid evidence reviews: impact and pathway to impact.

### 3.6 Learning from the WCEC knowledge mobilisation processes

A summary of what worked well, challenges and what could be improved is provided in Table 6.

**Table 6.** A reflection on enablers, challenges and lessons learnt.

| Area | Enablers, challenges and reflection |
| --- | --- |
| **Engagement and co-production with stakeholders** | • Working together with stakeholders from the outset is essential if research evidence is going to be relevant, timely and used by stakeholders.<br>• Co-production was essential to clarify the research question, identify priority outcomes and identify the policy and practice implications.<br>• The close working relationship with the Welsh Government's TAC and TAG ensured that the reviews were impactful e.g., directly informing Welsh Government policy officials in developing policy and plans.<br>• Close stakeholder involvement helped to build relationships and trust in the outputs of the reviews and the methods used.<br>• Stakeholders gained an understanding of the rapid review methods and limitations.<br>• Ensuring that stakeholders were able to attend and contribute to the stakeholder meetings could be challenging. This was mitigated by ensuring WCEC team flexibility and stakeholders sending representatives. |
| **Stakeholder informed knowledge mobilisation and accessibility of findings** | • Stakeholder input into the knowledge mobilisation plans was crucial to ensure findings reached relevant groups. They also provided considerable input to identify other groups that may be interested in the reports as part of the wider dissemination processes.<br>• Welsh Government evidence briefing sessions were useful to raise wider awareness of the findings, and to discuss policy and practice implications and knowledge mobilisation.<br>• Welsh Government evidence briefings were useful during the COVID-19 pandemic where stakeholders were meeting frequently as teams (such as TAC and TAG). However, moving out of the acute phase of the pandemic, it was more challenging to ensure attendance, and presenting at stakeholders' team meetings saw more attendees.<br>• Members of our public partnership group contributed equally as stakeholders and were able to fit in with the rapid timelines required. Their contribution was also invaluable to ensure the evidence was accessible to lay audiences. |
| **Ensuring accessibility of findings** | • Accessibility of findings is essential if stakeholders are to use the evidence. The key findings, implications and strength of the evidence were summarised within the 2-page Topline summary, which was well received.<br>• Infographics were useful but required additional time to produce. A clear need for the infographics should be evidenced.<br>• Even outputs made accessible to the general public (e.g., lay summaries, infographics, our on-line public events) may not be readily available to some people e.g. those who do not have access to electronic and digital media. Public-facing engagement events may mitigate this, and consideration of the additional time and resources needed to do this well should be planned.<br>• Other methods of increasing the accessibility of findings e.g., preparing videos to play in GP waiting rooms, producing booklets which could be picked up in pharmacies may be useful. Any approaches should ideally be accompanied by evaluation to assess effectiveness. |
| **Potential impact and pathway to impact** | • The research impact and pathway-to-impact and must be identified from the start.<br>• Meeting the timeframe in which the relevant research output is needed by stakeholders is essential for evidence to be used and impact realised.<br>• Tracking impact and use of information by Welsh Government advisors and decision-makers was occasionally challenging where the key stakeholders involved had moved to different departments or left. Having a Welsh Government member ‘boundary spanning’ between the WCEC and Welsh Government was invaluable to help identify new group leads and contact Welsh Government members to collect evidence of impact.<br>• Impact beyond the informing of a policy or plan i.e., actual patient and public impact can take years to be realised. Plans for long term follow up should be considered.<br>• Evidencing the impact of wider dissemination can be challenging. Asking people to enter details for further contact could help examine what numbers of downloads and visits to a site means.<br>• Social media highlighted the existence of the Centre and its work; however, evaluation did not include evidence of impact from social media. Tweets by Twitter users could be analysed for number, type of user, likes and dislikes to gauge reach and impact. |
| **Publication of reports** | • During the pandemic a delay of even 3 days would often not have been acceptable. It was essential to provide stakeholders with a watermarked (confidential) report prior to final editing and publication. Publishing in the WCEC library was immediate and could assist referencing in Welsh Government reports.<br>• Initially reports were hosted on a website and made accessible to view and download from our website library. However, if publishing in this way, reports do not have an individual identifier number (doi) so may not be picked by a Google search or search of published evidence resources. Additionally, when trying to publish an associated paper in a peer-reviewed journal the WCEC reports were identified by plagiarism software, which precluded the peer-reviewed publication despite clarifying this with journals.<br>• Publishing on pre-print servers was relatively rapid (3-7 days) and many journals accept publications previously available in this way. The reports also received a unique doi number and the preprint servers collected metrics including views, downloads, reference in policy documents and social media references i.e., in Twitter. |

## 4.0 Discussion

This paper describes the knowledge mobilisation processes of the WCEC, which were set up rapidly, and ensured that evidence was available to health and care decision-makers and advisers in Wales in response to the COVID-19 pandemic. Measures of success include that over 2 years, the WCEC worked closely with >30 stakeholder groups, including members of the public, to produce 51 evidence reviews, underpinned by knowledge mobilisation processes, which led to the evidence informing health and care policy and practice decisions during the pandemic.

Challenges, enablers, and lessons learnt in relation to knowledge mobilisation of evidence reviews have been included in Table 6. Some reflections on the five steps of our knowledge mobilisation framework are as follows.

### Step 1: Co-production and engagement with stakeholders

The successful engagement and co-production with a wide range of stakeholders in Wales may not have happened outside the urgency and unique requirements of the pandemic. The pandemic directly led to the set-up of the new Welsh Government TAC and TAG, who the WCEC was able to work with closely. Having a person that was embedded in both Welsh Government and the WCEC was crucial, and this has been evidenced elsewhere.(34) Other health and care groups also had an urgent need for evidence, which helped forge relationships and co-production.

The responsiveness and involvement of stakeholders was key to contextualise the review findings and expedite knowledge mobilisation. Involvement during the review process also built trust in the results and the WCEC processes. Moving beyond the pandemic, the need for evidence to address issues like long health service waiting lists, is ongoing, though the time frames may not have the same immediate urgency. A key challenge will be the maintaining the stakeholder relationships to ensure future research will contribute towards evidence informed policy and practice. Already structures and teams set up during the pandemic have changed. For example, the TAC and TAG have now dissolved, though their strength and contributions were realised, and a new Welsh Government Science, Evidence and Advice division has been set up, which includes members of TAC and TAG.

### Step 2: Stakeholder informed knowledge mobilisation

Involving stakeholders in knowledge mobilisation processes was crucial to enable evidence dissemination, and track use of the evidence. It enabled much wider sharing of evidence and links to be built with groups outside academia, which may not have been so readily accessible otherwise.

Knowledge mobilisation becomes particularly challenging where organisations are large and diverse and requires domain-specific strategies. Working with stakeholders to disseminate evidence ensures that evidence is seen as coming from a trusted source and reaches appropriate audiences. For example, the WCEC worked together with Social Care Wales knowledge exchange and research teams to disseminate evidence across social care organisations.

Reports of the evidence review findings, included the two-page Topline summary of headline findings. This was directly targeted at the needs of stakeholder and their feedback was positive. The Topline summary was based on other evidence policy briefs including the BRIDGE Evidence-informed framework for effective information-packaging to support policymaking.(7) Moving forward, it may be useful to include economic considerations, and qualitative research capturing public/patient experiences alongside results of evidence synthesis.(1)

In addition to tailored and targeted messages, training may increase evidence uptake by health managers and policymakers.(35)

### Step 3: Wider dissemination

Wider dissemination processes increased access to our reports and potentially avoided duplication of work by other Centres reviewing pandemic related evidence. It is worth noting that these processes required a considerable amount of time (e.g., developing and maintaining website and library, editing and publishing of reports, drawing up wider knowledge mobilisation plans, organising public events). Further evaluation to understand the meaning of metrics, such as downloads and social media mentions would have been useful. It is recognised that while passive knowledge dissemination increases access to evidence, it may not have any effect on uptake.(35) Future planning of wider dissemination with robust outcome measures and evaluation is needed.

It is worth noting that holding breakout sessions within one of the online public symposia worked well to engage wider public and identify their questions and priorities. Participants engaged in the follow-up processes including ranking their top 10 questions, which were included in the WCEC question prioritisation process.

### Step 4: Associated activities

Involving students in the work of the WCEC was valuable for both the WCEC and students. The students gained experience not only of conducting systematic reviews but also experience of knowledge mobilisation and of working with and providing evidence that informed stakeholders. Both the reviews and infographics produced by students were a valuable addition to the Centre outputs. The more in-depth requirements for academic student projects may not suit evidence synthesis where evidence need is very urgent as projects typically take longer to complete.

Identifying further research needs and gaps in evidence reviews should be valuable for researchers and to inform research funders. The funding organisations have specific requirements for considering such further research and evidence priorities, which need to be addressed.

### Step 5. Track and evidence impact

The best evidence of impact was for questions that came from the Welsh Government TAC and TAG groups, where the evidence was required to inform a specific plan, programme or guidance, and where knowledge mobilisation and gathering impact evidence was supported by stakeholders from the outset. While impact could be evidenced to the point of informing health and social care decision-makers (a recognised impact outcome)(13), evidence of how the research benefited public and patients would require future tracking and data analysis, which was not achievable within the timeline of the WCEC (2021-23). The unique environment of the pandemic and the close collaboration helped outputs to be used to inform decisions. However, moving forward, gaining a better understanding of barriers and facilitators to evidence use, how policymakers process evidence, and how other factors influence their decisions, will be crucial.(12,36)

Closer ‘knowledge brokering’ and embedding people to increase cross-community interactions may increase utilisation of evidence and impact.(34) Trade-offs between the resources required to enable this and increase in impact would be interesting to evaluate.

### The framework for our knowledge mobilisation processes

The ‘Knowledge to Action’ (KTA) framework was used to inform our model and processes (Fig. 1), as the stages aligned to many of our intended processes, and the model could be adapted to our rapid requirements. (Figure 1). The original KTA framework includes a knowledge creation process and a seven phase action cycle including 1) identifying the problem or issue to change, selecting knowledge to address the issue, determining the gap between knowledge and practice, 2) adapting the knowledge to your context, 3) assessing barriers and facilitators to knowledge use, 4) selecting an implementation strategy to make changes, 5) monitoring knowledge use, 6) evaluating outcomes and 7) sustaining the change or use of knowledge.(4) The time constraints and volume of evidence reports limited our implementation of some of these actions. However, our experiences and learning (Table 7), in addition to reflection on other models of knowledge mobilisation and communication will allow us to evolve our processes as we move forward.(35)

**Table 7.** Key learning points.

|  |
| --- |
| It is possible to rapidly set up a new evidence unit that can provide timely evidence to policy and practice decision making. This was facilitated by the urgency of the pandemic, but the model and lessons learnt are valuable to inform future initiatives. |
| To ensure that research findings feed into urgent decision-making processes, the timelines, outcomes, outcomes and the potential impact should be identified and agreed with the stakeholders from the outset. |
| Having a boundary spanning person, e.g., between the WCEC and Welsh Government, may be essential to enable good collaboration with Welsh Government groups, for sharing findings, and for evidencing and tracking impact. |
| Co-production with stakeholders and meeting their timelines was essential to identify policy and practice implications, enable knowledge mobilisation, and ensure evidence fed into urgent decision-making processes. |
| Ensuring stakeholders were involved during the evidence synthesis process (e.g., stakeholder and review team meeting) enabled an understanding of the methodology, its limitations and trust in the evidence outputs. |
| Involvement and contribution of public members was possible within all the WCEC knowledge mobilisation processes despite the rapid nature of the work. |
| Knowledge mobilisation processes should be flexible to meet stakeholder need. Knowledge mobilisation is a 2-way process and should underpin the work from start to finish - we can push the evidence out, but key stakeholders need to pull it into their practice. |

### Strengths and Limitations

Some of the strengths and limitations are described for each of the knowledge mobilisation steps above and in Table 6.

Strengths included the close involvement and co-production with stakeholders, including public partnership members. This ensured that evidence outputs were targeted, timely and used.

The WCEC survey to better understand use and impact of the evidence reviews is a limitation and while providing some insight, could not contribute to robust evaluation.

A further limitation warranting wider debate is what is recognised as impact. While rapid reviews could count as impact for informing policy and practice, the impact requirements for academia need to be further considered. Academia necessitates the publication of research in peer reviewed journals for both career development, and to underpin impact cases included in the UK Research Excellence Framework (which evaluates research impact of British Higher Education Institutions). While it is possible to further develop the rapid reviews in line with requirements of peer reviewed journals, this was only possible for 2 reviews during the lifetime of the WCEC, primarily owing to time constraints.

Lessons that will be taken forward include i) refining our engagement and communication with policy and practice decision makers, ii) developing a more robust and longer term knowledge mobilisation and evaluation plan from the outset, and iii) planning activities to better include and engage members of the public especially from under-served communities and those digitally excluded in Wales. Getting a public voice and stories of personal experiences would likely be a powerful addition to the evidence.(5) This should be feasible when not having to meet the quantity and urgency of work that needed to be undertaken during the public health emergency of the pandemic.

## Conclusion

The WCEC demonstrated that an Evidence Centre could be set up within a rapidly moving pandemic, with knowledge mobilisation processes ensuring that evidence reviews successfully informed health and social care decision-makers.

The value of the WCEC to inform evidence based decisions and practice has been recognised with a further five years of funding (from April 2023) as the ‘Health and Care Research Wales Evidence Centre’.

## Data Availability

All data produced in the present study are available upon reasonable request to the authors

## Acknowledgements

We would like to acknowledge the WCEC research partners from the Wales Centre For Evidence Based Care, the Specialist Unit for Research Evaluation, the Public Health Wales Evidence Service, Health Technology Wales and the Bangor Institute for Health and Medical Research, the BioComposites Centre, and the Cedar Health Technology Research Service for conducting the evidence reviews and presenting findings at stakeholder meetings including the Welsh Government evidence briefings and WCEC public symposia. We would like to thank our students Tomi Adewole and Kismet Lalli for their work, and also Peter Bragge, Lauren Elston and Sophie Hughes for their help in designing the stakeholder survey.

We would also like to acknowledge the members of our Public Partnership Group (PPG) and Barbara Harrington and Julie Hepburn from the Wales Primary and Emergency Care (PRIME) Service Users (SUPER) group for writing the lay summaries. Also, to members of the PPG for attending meetings with stakeholders and evidence briefings, Deb Smith for supporting our public engagement event, and the Welsh Government TAG secretariat in arranging evidence briefings.

We would like to thank all our stakeholders for giving their time and expertise and participating in knowledge mobilisation.

Lastly, we would like to thank the Health and Care Research Wales communications team for their support of the WCEC.

